# Effectiveness of four oral antifungal drugs (fluconazole, griseofulvin, itraconazole, terbinafine) in current epidemic of altered dermatophytosis in India: A randomized pragmatic trial

**DOI:** 10.1101/19003541

**Authors:** Sanjay Singh, Usha Chandra, Priyanka Verma, Vinayak N Anchan, Ragini Tilak

**Affiliations:** Department of Dermatology and Venereology, Banaras Hindu University, Varanasi, India; Department of Microbiology, Institute of Medical Sciences, Banaras Hindu University, Varanasi, India

**Keywords:** Dermatophytes, Dermatophytosis, Fluconazole, Griseofulvin, Itraconazole, Terbinafine, Tinea

## Abstract

**Background:** Dermatophyte infections have undergone unprecedented changes in India in recent past. Clinical trials comparing effectiveness of 4 main oral antifungal drugs are not available. We tested effectiveness of oral fluconazole, griseofulvin, itraconazole and terbinafine in chronic and chronic-relapsing tinea corporis, tinea cruris and tinea faciei.

**Methods:** Two hundred microscopy confirmed patients were allocated to 4 groups, fluconazole (5mg/kg/day), griseofulvin (10 mg/kg/day), itraconazole (5mg/kg/day), and terbinafine (7.5mg/kg/day), by concealed block randomization and treated for 8 weeks or cure. Effectiveness was calculated based on intention to treat analysis.

**Results:** At 4 weeks, 4, 1, 2, and 4 patients were cured with fluconazole, griseofulvin, itraconazole and terbinafine, respectively (P=0.417). At 8 weeks, 21 (42%), 7 (14%), 33 (66%) and 14 (28%) patients were cured, respectively (P=0.000); itraconazole was superior to fluconazole, griseofulvin and terbinafine (P≤0.016). Relapse rates after 4 and 8 weeks of cure in different groups were similar. Numbers-needed-to-treat (NNT) (versus griseofulvin), calculated based on cure rates at 8 weeks, for itraconazole, fluconazole, and terbinafine were 2, 4 and 8, respectively.

**Conclusion:** In view of cure rates and NNT, itraconazole is the most effective drug, followed by fluconazole (daily), terbinafine and then griseofulvin, in chronic and chronic-relapsing dermatophytosis in India.

**One Sentence Summary:** Effectiveness of all four antifungals has declined, with itraconazole being the most effective currently in dermatophytosis in India.

## Introduction

Unprecedented changes in the epidemiology, clinical features and treatment responsiveness of dermatophyte infections have been noted in recent past in India, with many patients presenting with chronic and chronic relapsing tinea.^1-4^ Recent data show that terbinafine, once a highly effective drug, now has an abysmal cure rate of 30.6% in tinea corporis, tinea cruris and tinea faciei when given orally at a dose of 5 mg/kg/day for 4 weeks.^5^ Decreased effectiveness of terbinafine has been attributed to mutation in the squalene epoxidase gene.^6, 7^ In the present study, we compared the effectiveness of four oral antifungal drugs, fluconazole, griseofulvin, itraconazole and terbinafine, for the treatment of chronic and chronic relapsing tinea corporis, tinea cruris and tinea faciei by performing a pragmatic, open, randomized controlled trial.

## Materials and Methods

### Setting

The study (registered with Clinical Trials Registry-India, registration number CTRI/2017/04/008281) was conducted at a tertiary care centre after obtaining approval from the Institutional Ethics Committee.

### Sample size

A sample size of 35 patients in each group was calculated considering the cure rates of 30% and 70%, with type I error rate of 0.05, type II error rate of 0.1 and dropout rate of 20%.^8^

### Selection criteria and enrolment

Patients with tinea corporis, tinea cruris or tinea faciei or any combination of these conditions were included in the trial. Inclusion criteria were (a) clinical suspicion of tinea corporis, tinea cruris or tinea faciei, (b) microscopic confirmation of tinea (potassium hydroxide [KOH] microscopy), (c) age between 4 years and 80 years, and (d) chronic and chronic-relapsing tinea (duration of tinea 3 months or more or tinea which was present for 3 months or more, got apparently improved within this period, but relapsed within a month).^9^ All these criteria had to be met for inclusion of a patient in the trial. Exclusion criteria included (a) pregnancy, (b) lactation, (c) presence of any other type(s) of tinea, e.g., onychomycosis, (d) history of drug reaction or allergy to any of the four drugs, (e) inability to come for follow-up every 2 weeks, (f) presence of any other disease requiring systemic therapy, and (g) abnormal results in complete blood count (CBC), liver function test (LFT), renal function test (RFT), plasma glucose (fasting and 2 hour post prandial test), glycosylated haemoglobin (HbA1C), fasting lipid profile (FLP), urine routine and microscopy examination (urine R/M), and electrocardiogram (ECG). Presence of any of the above exclusion criteria made a patient ineligible for the trial. A witnessed, written and informed consent was given by the patients, or by a parent in case of minor patients. All female patients of child bearing age were advised to avoid pregnancy during treatment.

Two hundred patients, who fulfilled the selection criteria, were randomly allocated to 4 parallel treatment groups on the basis of random numbers generated online using Research Randomizer (https://www.randomizer.org/). Random sequence was generated by block randomization and each group had 50 patients (allocation ratio 1:1:1:1). Allocation was concealed using sequentially numbered, sealed, opaque envelopes, which contained the allocation code written on a folded paper, the envelopes were opened after enrolment of the patients. Both random sequence generation and allocation concealment were done by a person unrelated to the trial. Patients were enrolled from April 2017 to March 2018. Skin scrapings taken at enrolment were inoculated on Sabouraud Dextrose Agar (SDA) with cycloheximide (0.05 g/l) and chloramphenicol (0.005 g/l). Identification of the dermatophyte was done based on colony morphology and microscopic morphology.

### Treatment

Doses given to the patients were as follows: fluconazole (5 mg/kg/day), griseofulvin (10 mg/kg/day), itraconazole (5 mg/kg/day), and terbinafine (7.5 mg/kg/day). Patients also received antihistamines (levocetirizine 5 mg in morning and hydroxyzine 25 mg at night and corresponding lower doses in children). Patients were asked to bring used strips of medicines and those in use at each visit. They were reminded to take the medicines as prescribed at each visit and received the treatment for 8 weeks or cure, whichever occurred earlier. Patients were advised against using any other treatment.

### Follow-up

Following data were recorded for each patient: body surface area (BSA) affected, treatment given, microscopy results, investigations at baseline and subsequent follow-ups, and follow-up data. The most severe lesion was identified as the index lesion, from which scraping and KOH microscopy was performed at baseline and then at follow-up visits.

Investigations performed at baseline in all groups were as follows: blood pressure measurement, CBC, LFT, RFT, plasma glucose fasting and post-prandial, HbA1C, ECG, FLP, urine R/M, blood pressure. During follow-up visits following investigations were performed: Fluconazole (CBC, RFT, FLP, ECG 4, 8 weeks; LFT 2, 4, 6, 8 weeks), Griseofulvin (CBC, LFT 2, 4, 6, 8 weeks; RFT, urine R/M 4,8 weeks), Itraconazole (CBC, RFT, FLP, urine R/M, ECG 4, 8 weeks; LFT 2, 4, 6, 8 weeks), and Terbinafine (CBC, LFT 2, 4, 6, 8 weeks).Patients were followed up at 2 weekly intervals up to a maximum of 8 weeks or cure, whichever occurred earlier. Patients who were not cured were treated outside the trial as follows: patient who were on itraconazole were prescribed terbinafine and patients on terbinafine, fluconazole, or griseofulvin were prescribed itraconazole.

### Measurement of treatment effect

Cure was defined as occurrence of both clinical cure (complete clearance of lesions) and mycological cure (negative KOH microscopy). Presence of post-inflammatory hyperpigmentation at the site of healed tinea was not considered a feature of tinea. Any of the following was considered treatment failure: (a) no improvement or worsening of the disease in 4 weeks in patient’s assessment, (b) presence of scaling and/or erythema at 8 weeks, and (c) positive microscopy at 8 weeks. Patients who were cured were asked whether they were fully satisfied with the treatment outcome or not.

### Outcome measures

Number of patients cured at 8 weeks with the different regimens was the primary outcome measure. Secondary outcome measures included number of patients cured at 4 weeks, relapse rate, and frequency of adverse events.

### Assessment for relapse

Those patients who were cured were assessed again 4 and 8 weeks after the cure to look for relapse of tinea (reappearance of erythema and scaling at the site of healed tinea).

### Statistical analysis

For the baseline variables, mean and standard deviation (SD) or median and range were calculated depending on the distribution of data. Cure rates and relapse rates were compared by Fisher’s exact test or Chi-square test, as appropriate. Cure rates with different drugs were calculated on the basis of intention-to-treat (ITT) analysis. P values less than 0.05 were considered significant. All P vales are two-tailed. Absolute risk reduction (ARR) of treatment failure at 8 weeks with fluconazole, itraconazole and terbinafine (versus griseofulvin) and number needed to treat (NNT) were calculated. When appropriate, 95% confidence intervals (CI) were calculated. Denominator for calculation of relapse rate was the number of patients who had achieved cure, those cured patients who could not be followed up were considered relapsed.

## Results

### Baseline characterises of the patients

Patients in the 4 groups were comparable at baseline with regard to age, weight, duration of illness and body surface area (BSA) affected [Table 1]. Out of 200 patients, 158 were male and 42 were female. Majority of the patients (98, 49 %) had tinea cruris et corporis, followed by tinea cruris et corporis et faciei (57, 28.5%), tinea cruris (28, 14%), tinea corporis (8, 4%), tinea cruris et faciei (3, 1.5%), tinea corporis et faciei (4, 2%) and tinea faciei (2, 1%). Cultures for dermatophytes were attempted in 83 patients, of whom dermatophytes grew in 28 cases. In all these cases, the dermatophyte was Trichophyton mentagrophytes.

**Table 1.** Baseline characteristics of the patients.

|  | Fluconazole<br>n=50 | Griseofulvin<br>n=50 | Itraconazole<br>n=50 | Terbinafine<br>n=50 |
| --- | --- | --- | --- | --- |
| Age (years)<br>(mean±SD) | 30.5 ± 13.6 | 28.2 ± 11.6 | 27.2 ± 10.1 | 31.1 ± 14.5 |
| Female patients, n (%) | 8 (4) | 10 (5) | 13 (6.5) | 11 (5.5) |
| Duration (months),<br>median (range) | 6 (3-36) | 5.5 (3-36) | 6 (3-72) | 6 (3-36) |
| Weight (kilograms)<br>(mean±SD) | 66.5±15.1 | 60.4±11.4 | 62.5±12.9 | 63.9±15.1 |
| Body surface area<br>(BSA) affected*<br>(mean±SD) | 0.7±0.2 | 0.7±0.2 | 0.7±0.2 | 0.7±0.3 |

\*Affected BSA was semi-quantitatively assessed on a 0 to 6 scale following the area grading method of Psoriasis Area and Severity Index.

### Cure rates at 4 weeks

In the fluconazole, griseofulvin, itraconazole and terbinafine groups, 4, 1, 2 and 4 patients were cured, respectively. Cure rates were not significantly different (P=0.417, Fisher’s exact test) [Table 2, Figure 1] (intention to treat analysis).

**Table 2.** Number of patients cured at 4 and 8 weeks with the four treatments.

| Treatment | 4 weeks* |  | 8 weeks** |  |
| --- | --- | --- | --- | --- |
|  | Number cured<br>(%), ITT | 95 % CI (%) | Number cured<br>(%), ITT | 95 % CI (%) |
| Fluconazole | 4 (8) | 2.6 to 19.4 | 21 (42) | 29.4 to 55.8 |
| Griseofulvin | 1 (2) | <0.01 to 11.5 | 7 (14) | 6.6 to 26.5 |
| Itraconazole | 2 (4) | 0.3 to 14.2 | 33 (66) | 52.1 to 77.6 |
| Terbinafine | 4 (8) | 2.6 to 19.4 | 14 (28) | 17.4 to 41.8 |

CI, confidence interval; ITT, intention-to-treat analysis. \*P=0.417 (Fisher's exact test),
\*\*\*P=0.000 (Chi-square test).

**Figure 1.**
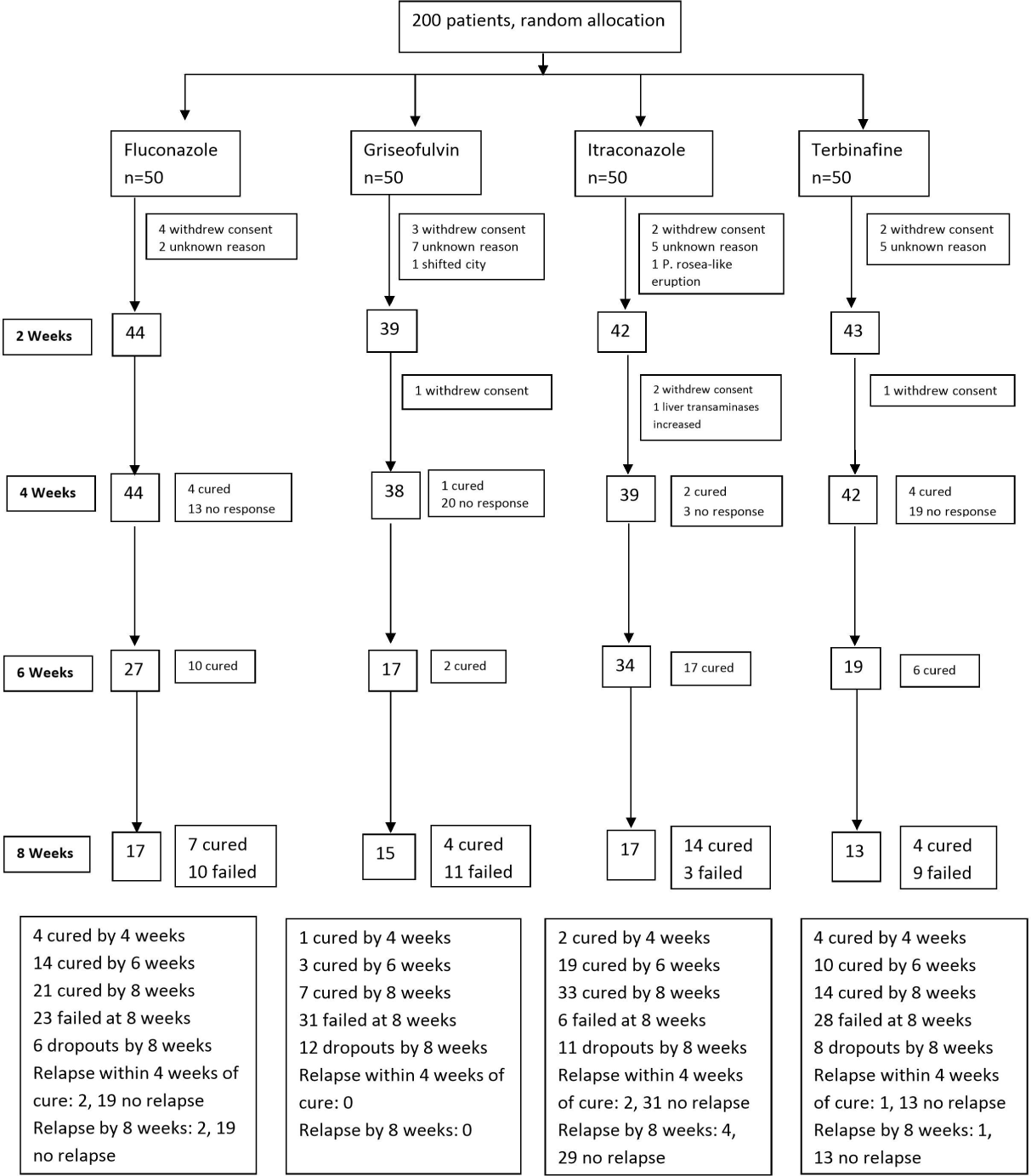
Flow diagram of the study; P. rosea, Pityriasis rosea.

### Cure rates at 8 weeks

In the fluconazole, griseofulvin, itraconazole and terbinafine groups, 21, 7, 33 and 14 patients were cured respectively [Table 2, Figure 1]. There numbers include the number of patients who were cured at 4 weeks. At least one pair of groups was significantly different for cure rates at 8 weeks (P=0.000, Chi-square test) (intention to treat analysis).

Six different pairs of the treatment groups were compared for cure rates at 8 weeks [Table 3]. Itraconazole was more effective than fluconazole, griseofulvin and terbinafine. Fluconazole was more effective than griseofulvin. Fluconazole and terbinafine, and griseofulvin and terbinafine had similar effectiveness. All cured patients were fully satisfied with the outcome.

**Table 3.** Comparison of cure rates at 8 weeks with the four treatments.

|  | P* | 95% confidence interval of difference |
| --- | --- | --- |
| Fluconazole vs. griseofulvin | 0.0018 | 10.5 to 43.5 |
| Fluconazole vs. itraconazole | 0.0161 | 4.5 to 41.1 |
| Fluconazole vs. terbinafine | 0.1422 | -4.6 to 31.3 |
| Griseofulvin vs. itraconazole | 0.0001 | 33.6 to 65.5 |
| Griseofulvin vs. terbinafine | 0.0857 | -2.1 to 29.4 |
| Itraconazole vs. terbinafine | 0.0001 | 18.6 to 53.6 |

\*Chi-square test.

### Number needed to treat

Numbers needed to treat (NNT) (versus griseofulvin), calculated on the basis of cure rates at 8 weeks, for itraconazole, fluconazole and terbinafine were 2, 4 and 8, respectively [Table 4].

**Table 4.** Number needed to treat and absolute risk reduction with fluconazole, itraconazole and terbinafine versus griseofulvin.

| **Treatment** | **NNT** | **ARR, %** | **95% CI of ARR, %** |
| --- | --- | --- | --- |
| Fluconazole | 4 | 28.0 | 11.3 to 44.7 |
| Itraconazole | 2 | 52.0 | 35.7 to 68.3 |
| Terbinafine | 8 | 14.0 | -1.7 to 29.7* |

Absolute risk reduction of treatment failure versus griseofulvin.
ARR, absolute risk reduction; CI, confidence interval; NNT, number needed to treat.
\*As the 95% CI of ARR extends from a negative number to a positive number, terbinafine may or may not be superior to griseofulvin.

### Adverse effects

Adverse effects were noted only in the itraconazole group in 2 male patients, in both the treatment was stopped. One patient developed pityriasis rosea-like rash on day 13 of treatment, the rash resolved in 2 weeks after stopping treatment. Another patient had asymptomatic increase in hepatic transaminases (serum glutamic oxaloacetic transaminase 68 units/litre, serum glutamic pyruvic transaminase 127 units/litre) at 4 weeks. He was seen 2 weeks later, when he was clinically normal, but he did not repeat liver function tests. Patients in the other groups experienced no adverse effects and none were detected on investigations.

### Relapse rates

At 4 weeks, 2, 0, 2, and 1 patients relapsed in fluconazole, griseofulvin, itraconazole, and terbinafine groups, respectively (P=0.420, Fisher’s exact test). Relapse rates after 8 weeks of cure in different groups were also not significantly different (P= 1.0, Fisher’s exact test) [Table 5, Figure 1].

**Table 5.** Relapse rates 8 weeks after cure with the four treatments*.

|  | Fluconazole | Griseofulvin | Itraconazole | Terbinafine |
| --- | --- | --- | --- | --- |
| Number of patients cured | 21 | 7 | 33 | 14 |
| Number of patients followed up | 20 | 7 | 32 | 13 |
| Number of patients with relapse | 1 | 0 | 3 | 0 |
| Number of patients not followed up | 1 | 0 | 1 | 1 |
| Total number of patients relapsed (%) | 2 (9.5) | 0 | 4 (12.1) | 1 (7.1) |

\*P= 1.0 (Fisher's exact test).

## Discussion

Present study was conducted in view of unprecedented changes in the epidemiology, clinical features and treatment responsiveness of tinea infections in recent past in India.^1-4^ This has led to an epidemic of altered dermatophytosis characterized by chronic and chronic-relapsing cases.^9^ Recent data show that terbinafine, once a highly effective drug, now has abysmal cure rate in tinea corporis, tinea cruris and tinea faciei.^5^ These results confirmed the general impression among Indian dermatologists that terbinafine is fast losing its effectiveness. There is also a general impression that effectiveness of other antifungal drugs has also declined, however, there is no published data on the effectiveness of oral antifungal drugs in dermatophytosis performed during the current epidemic in India.

We tested the effectiveness of oral fluconazole (5mg/kg/day), griseofulvin (10 mg/kg/day), itraconazole (5mg/kg/day) and terbinafine (7.5mg/kg/day) for the treatment of tinea corporis, tinea cruris and tinea faciei of 3 months or more. The trial was conducted within a pragmatic setting. Two hundred microscopy confirmed patients were assigned to the 4 treatments by concealed random allocation and treated for 8 weeks or cure, whichever occurred earlier.

At 4 weeks, the four treatments were similarly ineffective. At 8 weeks, 21, 7, 33 and 14 patients were cured with fluconazole, griseofulvin, itraconazole and terbinafine, respectively. Itraconazole was superior to fluconazole, griseofulvin and terbinafine, and fluconazole was superior to griseofulvin. Numbers needed to treat (NNT) (versus griseofulvin), calculated based on cure rates at 8 weeks, for itraconazole, fluconazole, and terbinafine were 2, 4 and 8, respectively, meaning thereby that, compared to griseofulvin, 2 patients need to be treated with itraconazole to achieve one additional cure, while 4 and 8 patients need to be treated with fluconazole and terbinafine for the same effect, respectively. Thus, itraconazole was the most effective treatment out of the four treatments tested in the study.

Relapse rates after 4 and 8 weeks of cure in different groups were not significantly different. Of the 200 patients, only 2, who received itraconazole, experienced adverse effects, one developed pityriasis rosea-like eruption and another had asymptomatic elevation of hepatic transaminases. However, the importance of closely monitoring liver enzymes cannot be overemphasized when treating patients with itraconazole.

The recently published study from India on patients with tinea corporis, tinea cruris and tinea faciei showed that cure rate with terbinafine (5 mg/kg/day) given for 4 weeks was 30.6%.^5^ In the present study, cure rates with terbinafine (7.5mg/kg/day) were 8% at 4 weeks and 28% at 8 weeks. These data suggest that effectiveness of oral terbinafine in tinea has declined further compared to the very recent data. However, this cannot be definitively commented upon because the present study was not designed to find out effectiveness of a single agent, which would require a larger sample size.

A limitation of the present study is that it was not blinded.

Clinical trial is the final arbiter on the effectiveness of a drug. In this pragmatic study, the objective was to compare the effectiveness of 4 commonly used oral antifungal drugs for tinea infections in a real-world setting, so that the results are relevant and applicable to how patients with tinea are treated in usual care. To conclude, in view of cure rates and NNT, itraconazole was the most effective drug for the treatment of tinea corporis, tinea cruris and tinea faciei, followed by fluconazole (daily), terbinafine and then griseofulvin in the current epidemic of dermatophytosis in India.

## Data Availability

Data not available online.

